# Global Longitudinal Active Strain Energy Density (GLASED): Age and Sex Differences in young and Veteran Athletes

**DOI:** 10.1101/2023.08.22.23294454

**Authors:** David H. MacIver, Henggui Zhang, Christopher Johnson, Efstathios Papatheodorou, Gemma Parry-Williams, Sanjay Sharma, David Oxborough

## Abstract

Graphic abstract

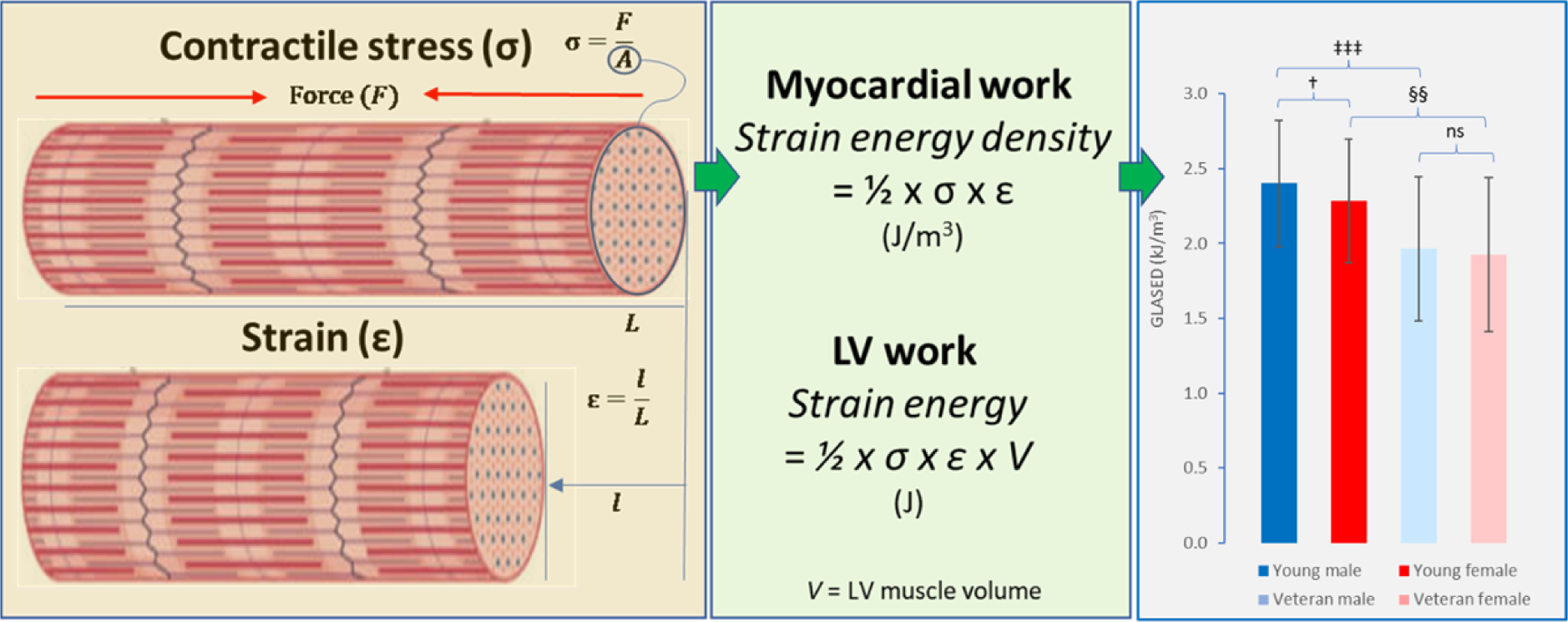

**Clinical perspective:** Global longitudinal active strain energy density (GLASED) is a recently introduced potential measure of ventricular function that combines myocardial stress and strain information. GLASED estimates the work performed per unit volume of myocardium during contraction. Recent studies with cardiac MRI have demonstrated that GLASED predicts prognosis more accurately than ejection fraction or strain alone. Our current study uses echocardiography and reveals previously unknown physiological differences in myocardial function between male and female athletes, as well as among young and veteran athletes. Our results suggest that GLASED could be a valuable tool in assessing cardiac diseases, particularly when the clinical phenotype is uncertain

**Background:** Global longitudinal active strain energy density (GLASED) is an innovative method for assessing myocardial function by quantifying the work performed by the left ventricular muscle. The use of GLASED holds promise for improving the diagnosis and management of cardiac diseases. This study aimed to evaluate the feasibility of measuring GLASED using echocardiography and investigate potential differences in GLASED values among athletes based on age and sex.

**Methods and Results:** An observational echocardiographic study was conducted, involving male controls, male and female young athletes, and male and female veteran athletes. GLASED was calculated from the myocardial stress and strain. The mean age (years) of young athletes was 21.6 for males and 21.4 for females, while the mean age of veteran athletes was 53.5 for males and 54.2 for females. GLASED was found to be highest in young male athletes (2.40 kJ/m^3^) and lowest in female veterans (1.96 kJ/m^3^). Veteran males exhibited lower values (1.96 kJ/m3) compared to young male athletes (P<0.001). Young females demonstrated greater GLASED (2.28 kJ/m^3^) than veteran females (P<0.01). However, no significant difference in GLASED was observed between male and female veterans.

**Conclusions:** Our findings demonstrate the feasibility of measuring GLASED using echocardiography. GLASED values were higher in young male athletes compared to female athletes, and it decreased with age. Importantly, the sex-related differences observed in GLASED values among young athletes were no longer present in veteran athletes. Estimating GLASED may serve as a valuable screening tool for cardiac diseases in athletes, particularly for those with borderline phenotypes of hypertrophic and dilated cardiomyopathies.

## Introduction

Long-term intense exercise induces changes in the left ventricles of athletes, including increases in wall thickness and ventricular volumes. These geometric changes are influenced by factors such as age, sex, training duration, sport type, and genetic factors.^1, 2^ Although the left ventricular ejection fraction (LVEF) has been the main measure of systolic function for over 50 years, recent studies have questioned the reliability of LVEF due to the impact of structural changes. Specifically, LVEF is increased by an increase in wall thickness^3, 4^ or a decreased internal diameter,^5^ and length^6^ independently of any change in myocardial strain. These modelling findings have recently been corroborated by other groups^7, 8^ and in clinical studies.^1, 6, 8^

The influence of each structural change on LVEF can be represented by parabolic curves described using quadratic functions (Appendix Section 1.). For example, a 1 mm increase in end-diastolic wall thickness (EDWT), the LVEF increases by between 2.1 and 2.6 percentage points. Conversely, an increase in left ventricular internal diameter in diastole (LVIDd) by 1 mm decreases LVEF by 0.4 to 1.2 percentage points. The corrected ejection fraction (EF_c_) was developed to account for the effects of geometric differences and expose the misleading nature of LVEF.^6^

Myocardial strain has been introduced to address the limitations of LVEF. Increasing myocardial strain magnitude increases LVEF with midwall circumferential strain having the greater contribution (2/3) compared with long-axis shortening (1/3).^9^ A change in midwall circumferential strain can alter LVEF by 2-3 percentage points for every 1% change in strain (Appendix Section 1.). Myocardial strain is often reduced in thicker walled ventricles, frequently with preservation of the LVEF ^10, 11^ due to a maintenance of absolute wall thickening.^6^ However, myocardial strain is notably affected by the afterload^12^ limiting its usefulness.

A new method for assessing contractile function called contractance or myocardial active strain energy density (MASED) estimates the mechanical work (energy) performed per unit volume of myocardial tissue.^12, 13^ MASED overcomes the weaknesses of both LVEF and myocardial strain by combining information from both the stress (contractile force per unit cross sectional area) and myocardial strain. MASED allows for loading conditions and can be applied to in vitro/ex vivo ^12^ and in vivo studies^13^ with an improved ability to compare research studies. In whole heart preparations and in vivo, MASED can be estimated in both the longitudinal and circumferential directions with the global longitudinal active strain energy density (GLASED) and circumferential active strain energy density (CASED) respectively.^13^

The work done or mechanical energy generated by muscle mass in the longitudinal and circumferential directions is called the global longitudinal active strain energy (GLASE) and circumferential active strain energy (CASE) respectively.^13^

We recently performed an assessment of GLASE, CASE, GLASED and CASED in cohorts with severe hypertension, dilated cardiomyopathy and amyloid heart disease using cardiac magnetic resonance imaging (CMR) and found that GLASED is the most accurate method for predicting the expected mortality in these conditions.^13^ GLASED is also more accurate than LVEF, corrected LVEF, strains, stresses, forces, stroke work, myocardial contraction fraction and pressure strain loops in predicting outcome in these diseases and is sensitive enough to detect changes in hypertensive cardiomyopathy (Appendix Section 2).^13^ Moreover, GLASED has the highest proportional hazard ratio for major adverse cardiovascular events and mortality when compared with strain, LVEF and all other potential structural and functional markers in a community-based cohort comprising 44,957 individuals using CMR (Appendix Section 3 and 4).^14^

In this study, we aimed to determine if GLASED could be assessed using echocardiography. Our pre-specified null-hypothesis was that there would be no difference in GLASED between the sexes and age groups. Hence, we sought to assess GLASED in young and veteran male and female athletes with a young male non-athlete control group to explored potential differences based on age and sex.

## Methods

### Cohorts

A retrospective analysis of 447 healthy individuals consisting of 5 cohorts, 245 young male athletes (mixed sports), 67 young female athletes (football/soccer), 70 veteran male athletes (mixed sports), 44 veteran female athletes (mixed sports) and 21 healthy non-athletes. Data from one of these cohorts has been published.^1^ Data were collected either as part of the mandatory pre-participation cardiac screening or as part of a planned and structured research study. Participants completed a health screening questionnaire to detail any cardiovascular symptoms, family history of sudden cardiac death or other cardiovascular history. Blood pressure was recorded using a standard sphygmomanometer. A resting 12-lead ECG and transthoracic echocardiogram was performed on all participants. A sports cardiologist reviewed all results. No individuals were excluded on imaging or clinical grounds. Ethics approval was obtained from the ethics committee of Liverpool John Moores University and St Georges University Hospital.

### Echocardiography

A standard echocardiogram was performed by British Society of Echocardiography (BSE) accredited experienced sonographers using a commercially available ultrasound system (Vivid Q or Vivid E95, GE Healthcare, Horten, Norway) with a 1.5-4 MHz phased array transducer, with the participant lying in the left lateral decubitus position. All images were attained in accordance with the BSE guidelines.^15^ Images were stored as a raw digital imaging and communications in medicine (DICOM) format and exported to an offline analysis system (EchoPac version 202, GE Healthcare, Horton, Norway) for subsequent analysis. The mean E’ velocities were calculated from the mean of the medial and lateral values using tissue Doppler. Doppler studies were not performed in the veteran male cohort.

The parasternal short-axis orientation was used to calculate end-diastolic wall thickness (EDWT) and left ventricular end-diastolic internal diameter (LVIDd) was obtained from the basal and mid anteroseptum, inferoseptum, inferior, posterior, lateral and anterior walls. Basal short-axis was located at the tip of the MV and mid short-axis at papillary muscle level. Each segment was measured once at each basal and mid-level in each of the 6 segments. The mean EDWT and mean LVIDd were calculated from the 12 wall thicknesses and 6 LVIDd dimensions.

Integrated myocardial speckle tracking software was used to calculate longitudinal and circumferential strain. Apical 4, 3 and 2 chamber orientations were used to derive global longitudinal strain (GLS) whilst parasternal short axis at basal, mid and apical levels were acquired for global circumferential strain. Images were optimised to maximise endocardial delineation and frame rates were maintained between 40-90 fps. Offline analysis allowed for semi-automated tracking and all images had acceptable tracking of all segments. Reproducibility for measuring strain has previously been reported as good in our laboratory using a repeated measures acquisition study.^16^ Circumferential strains were not available for the young female athletes. Left ventricular muscle mass was calculated using the Penn equation.^17^

### Active strain energy density

Peak longitudinal and midwall circumferential nominal stresses were calculated using the Lamé equations. The Lamé equations were used for calculating nominal stresses, as the Laplace method is only accurate for thin-walled chamber with a diameter/thickness <20,^13, 18^ as follows:

Longitudinal Lamé stress

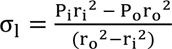

Midwall circumferential Lamé stress

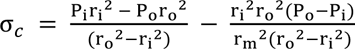

where P_i_ is inner (ventricular cavity) pressure (in Pa) and equal to peak systolic pressure. P_o_ is outer (pericardial) gauge pressure and is assumed to be 0 Pa.^19^ The brachial cuff derived from systolic blood pressure was used for P_i_. Further, r_m_ is midwall, r_o_ is outer (epicardial), and r_i_ is inner (luminal or endocardial) LV radii, respectively.

Nominal longitudinal force was calculated from the product of longitudinal Lamé stress and the end-diastolic short-axis cross sectional myocardial area.

GLASED and CASED were calculated using the following equations.^13^

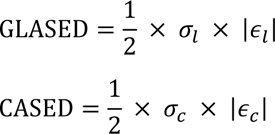

Where σ_l_ is the longitudinal nominal stress, σ_C_ is the midwall circumferential nominal stress, |ε_l_| is the magnitude (absolute value) of peak longitudinal strain and |ε_C_| is the magnitude of peak circumferential strain.

The sum of active strain energy densities (SASED) was calculated by adding GLASED to CASED.

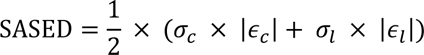

GLASE and CASE were calculated by multiplying GLASED and CASED respectively by LV muscle volume derived from the muscle mass by assuming a myocardial density of 1.05 g/ml.^20^

### Relationship between LVEF and structural differences

A modelling substudy was performed to assess the impact of differences in left ventricular geometry and strain on LVEF to facilitate understanding of the differences in LVEF found in our in the athletes. The method has been described in detail elsewhere.^1, 6^ The two-shell model was used and the left ventricular end-diastolic diameter, wall thickness and strain were altered to assess their effect on LVEF. Changes in LVEF were obtained by individually adjusting the input variables as follows: midwall circumferential shortening 15-20%, EDWT 10 to 15 mm and LVIDd 40-50 mm. Quadratic equations were derived from the resulting curves allowing the relative impact of each variable to be calculated (see Appendix 1.). The equations obtained were then used to calculate the expected differences in LVEF from the relevant input variables from the cohorts and compared with the measured LVEF.

### Statistical analysis

Our primary a priori null hypothesis was that there was no significant difference in GLASED between sexes in the different age groups. Normality was assessed using the Shapiro-Wilk test. Either a two tailed T-tests or Mann-Whitney tests were performed on these pairs as appropriate. To allow for multiple comparisons, a one-way analysis of variance (ANOVA) was performed for all comparisons with Tukey HSD/KRAMER analysis when both cohorts had a normal distribution and Kruskal-Wallis Test/Nemenyi when either cohort was not normally distributed. Correlations were performed using Pearson’s method.

## Results

### Demographic and echocardiographic findings

No individuals had clinically significant valvular disease or any overt cardiomyopathic processes. All results and statistical significances are shown in Table 1. There was no significant difference in age between young male and young female athletes (21.6 and 21.4 years respectively, ns). Veteran males and females had a similar age (53.5 vs 54.2 years, ns).

**Table 1:**
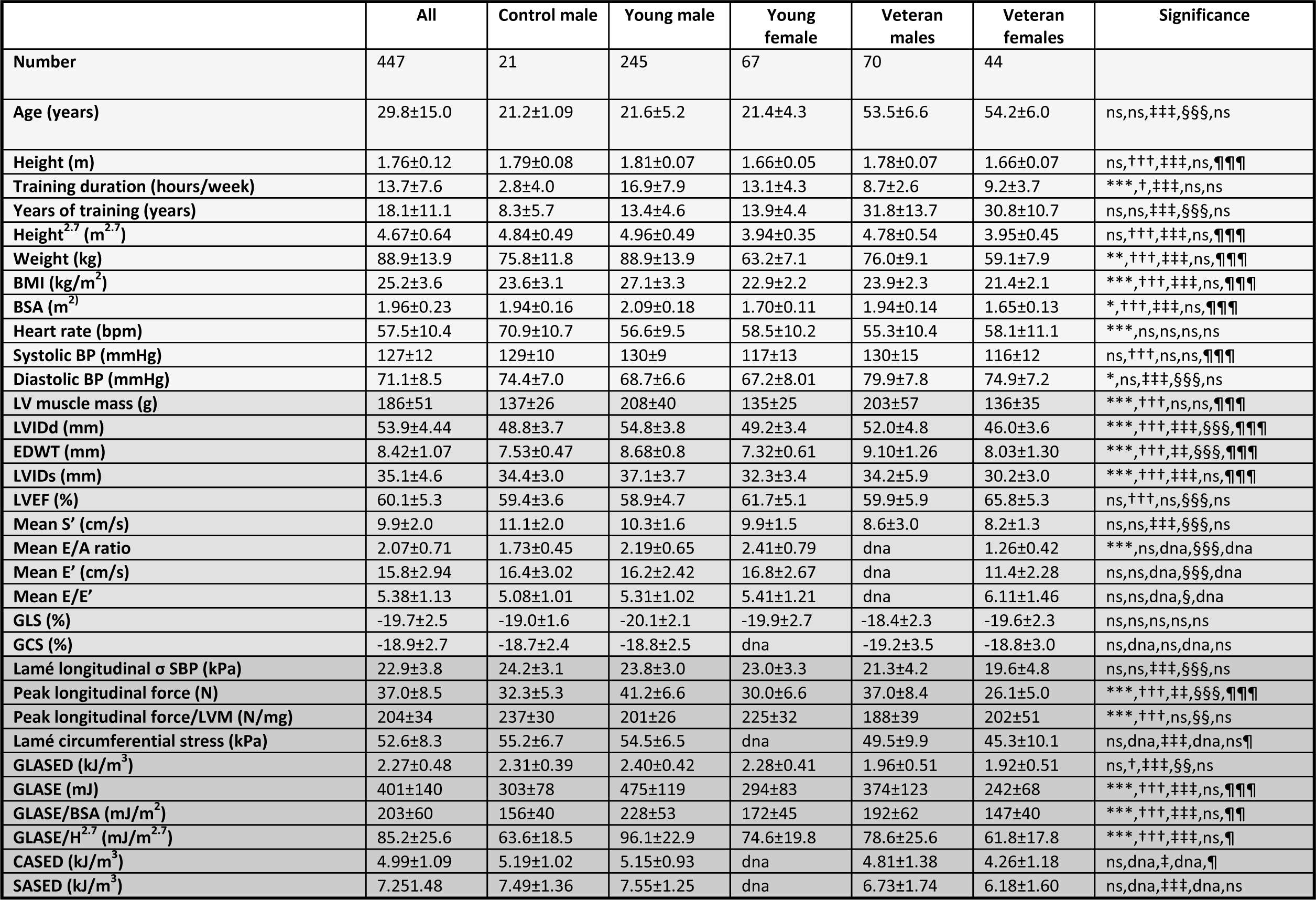
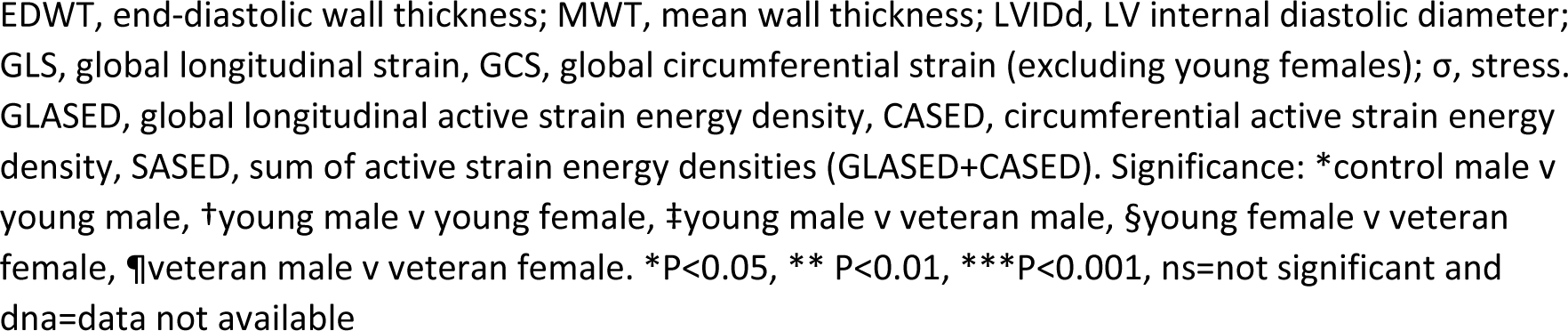
Demographics and results (mean±1SD)

Young male athletes were taller, heavier, had a greater BMI and BSA, higher SBP, LV mass, LVIDd, MWT and EDWT compared with young female athletes. Veteran males were taller and heavier than veteran females. There was no significant difference in heart rates between the athletes but control males had higher rates. Male athletes had greater systolic blood pressures compared with female athletes and veteran athletes had higher diastolic blood pressures compared to young athletes. respectively. LVEF was higher in females in both age groups with the difference in LVEF being greater in the veterans.

Mitral E/A ratio was higher in young males compared to young females and in young females vs veteran females. Mean E’ was the lowest and E/E’ the highest in veteran female athletes. S’ was lower in veteran athletes but there were no differences between the sexes (Table 1)

### Myocardial stresses and strains

Global longitudinal strain was not statistically different in males and females or young athletes and veterans (Figure 1A). Longitudinal contractile wall stress was higher in young male athletes compared with veteran athletes with the greatest decline in veteran females (Figure 1B). Midwall circumferential contractile stress was higher in young athletes compared with veteran athletes.

**Figure 1:**
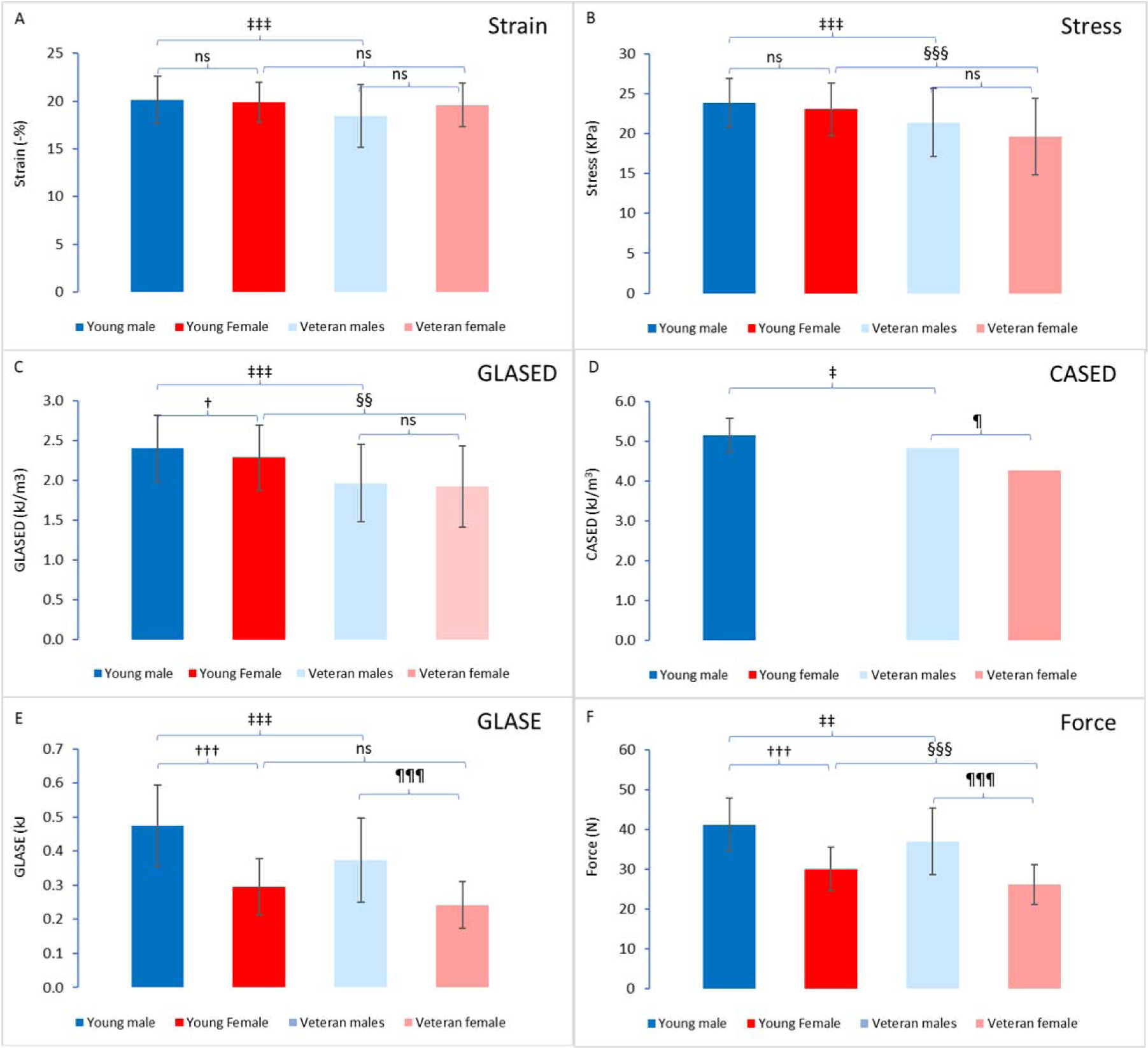
Graphs showing the values for longitudinal strain, longitudinal stress, GLASED, CASED, GLASE and peak longitudinal force.

### GLASED, CASED and SASED

GLASED was calculated in all individuals. GLASED was highest in young males athletes (2.40 kJ/m^3^) and significantly greater than young female athletes (2.28 kJ/m^3^, *P*<0.05) (Figure 1C). Male and female veterans had a significantly lower GLASED (1.96 and 1.92 kJ/m3, *P*<0.001 and P<0.01 respectively) compared with their sex matched cohorts (Figure 1C and Figure 2). Young male non-athletes had a trend to lower GLASED values (2.27 kJ/m^3^) compared to young male athletes (not significant). Circumferential contractance (CASED) was lower in veteran males compared with young athletes (Figure 1D). Female veterans had lower circumferential contractance compared with male veterans. CASED data for female athletes could not be calculated as circumferential strain data was not available.

**Figures 2:**
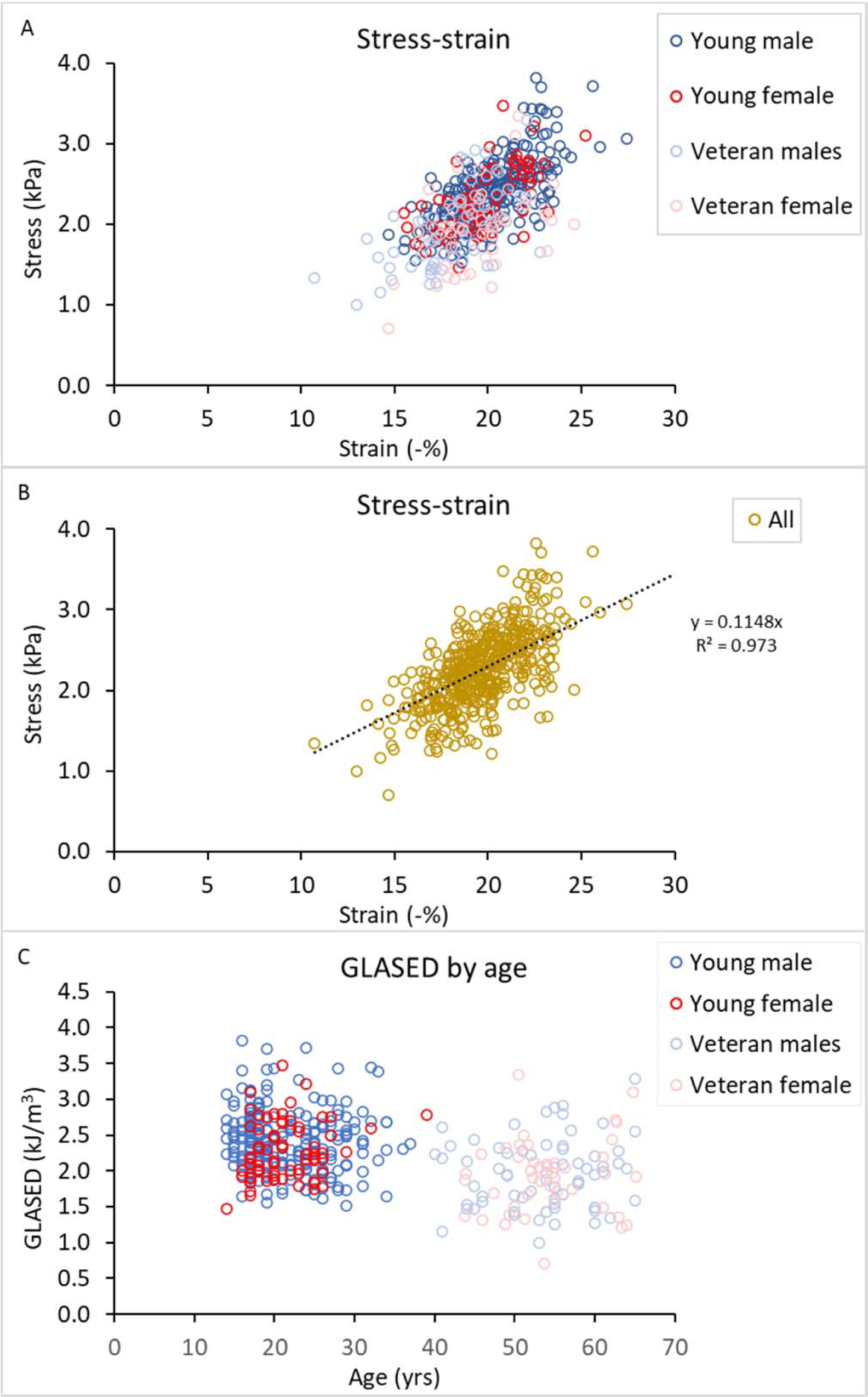
Scatter plots showing A. Relationship between stress and strain by cohort. B. Stress strain relationship for all cohorts and C. relationship between GLASED and age.

### GLASE and CASE

Left ventricular work was assessed using GLASE. GLASE was highest in the young male athletes and decreased in the veteran males (Figure 1E). Female athletes had lower GLASE compared with male counterparts. Young female athletes had similar GLASE values to veteran female athletes. GLASE was higher in young male athletes compared with young male non-athletes (475 vs 303 mJ, *P*<0.001)

### Longitudinal forces

Males had significantly greater longitudinal peak force compared with females and younger athletes had higher forces than older athletes (Figure 1F).

### Relationship between stress and strain

Figure 2A shows the relationship between stress and strain in each cohort. Myocardial strain magnitude increases (is more negative) as myocardial contractile stress increases. Figure 2B shows the same data for all cohorts including male controls and shows a good correlation with R^2^ of 0.931 when intercept is set at 0,0 and 0.64 when intercept was not pre-defined (*P*<0.0001).

### Relationship between GLASED and age

Figure 2C shows scatter plots of the association of age with GLASED showing lower values in the older age groups.

### Relationship between LVEF and structural differences

The modelling substudy confirmed that changes in LVEF are curvilinear with each individual variable with steepness and shape of slope dependent on each of the other variables (see Appendix Section 1. for further details). These figures represent the changes based on an alteration of a single variable with the other variables fixed as follows: LVIDd 45 mm, EDWT 10 mm and magnitude of midwall circumferential shortening of 18.7%.

The substudy showed that the higher LVEF observed in female athletes compared to their male counterpart can be explained by the differences in structure, namely the lower end-diastolic diameter despite a lower EDWT (Table 2.).

**Table 2:** Change in LVEF by structural differences and strain.

| **Variable** | **LVEF** | **Regression coefficient\*** | **Mathematical model\*\*** | **References** |
| --- | --- | --- | --- | --- |
| ↑GLS | ↑↑ | +0.65/% | - | 6, 9 28 |
| ↑mwCS | ↑↑↑ | +2.1/% | +3.2/% | 6, 9 28 |
| ↑EDWT | ↑↑↑ | +2.1/mm | +2.6/mm | 3 6 28 |
| ↑LVIDd | ↓↓ | -0.42/mm | -1.2/mm | 6, 28 |
| ↑LV length | ↓ | -0.17/mm | - | 6 |

GLS, magnitude (absolute value) of global longitudinal strain; mwCS, magnitude of midwall circumferential strain; EDWT, end-diastolic wall thickness; LVIDd, left ventricular internal diameter in diastole; LV length, left ventricular internal long axis length in diastole.
\*Obtained using five variable linear statistical model.
\*\*Derived from 5 variable mathematical model used in<sup>6</sup>.

**Table 3:** Reference ranges for GLASED in the different cohorts– assumes ±1.96×1SD (age range of cohorts).

| **GLASED**<br>**(kJ/m<sup>3</sup>)** | **All**<br>**(14-65)** | **Male**<br>**control**<br>**(20-25)** | **Male**<br>**athlete**<br>**(14-37)** | **Female**<br>**athlete**<br>**(14-39)** | **Male**<br>**veteran**<br>**(40-65)** | **Female**<br>**veteran**<br>**(44-65)** |
| --- | --- | --- | --- | --- | --- | --- |
| **Upper** | 3.21 | 3.08 | 3.22 | 3.09 | 2.91 | 2.93 |
| **Lower** | 1.34 | 1.54 | 1.58 | 1.48 | 1.02 | 0.92 |

## Discussion

To the best of our knowledge this is the first echocardiographic study to assess left ventricular contractance. The study confirmed that estimating longitudinal contractance by calculating GLASED using echocardiography was practical and can be readily implemented in clinical practice.

Athletes are known to have altered left ventricular geometry^2^ that impact traditional measures of myocardial systolic function such as LVEF.^1^ Myocardial strain is influenced by afterload^12^ and there are differences in blood pressure between the sexes in athletes.^21^ Strain energy density is calculated from stress (i.e. wall thickness, diameter and pressure) and strain and has a long standing background in engineering science. GLASED provides theoretical advantages over established methods as GLASED corrects for potential confounders such as differences in afterload (e.g. blood pressure) and ventricular remodelling.

We showed significant a significantly higher contractance in young males compared to young females and young athletes compared to veteran athletes. Potential alternative measures of cardiac function such as myocardial strain and LVEF in isolation were unhelpful in distinguishing between the sexes. Plausible explanations for the sex differences in GLASED include intrinsic genetic or hormonal differences, type of sports undertaken, training methods and levels of fitness.

The higher GLASED in younger athletes compared with veterans suggest that they can produce more mechanical work (energy) per unit volume of myocardium and may indicate that the cardiac muscle is intrinsically stronger for a given unit mass. Non-specific age-related deterioration, type of sport or training regime, prolonged training damage with myocardial cell death and replacement fibrosis, or hormonal changes could be explanations for the lower values of contractance in the veteran cohorts.

The calculation for LVEF is obtained from ventricular luminal information alone and yet is influenced by changes in geometry and strain. It has previously shown that a larger internal diameter^5^ and length decreases LVEF^6^ whereas a greater EDWT increases LVEF.^1, 3^ The combination of higher LVIDd and EDWT has opposing effects on LVEF.^3, 5^ Previous studies have also shown a higher LVEF in females compared to males.^1, 22, 23^ Our finding of higher LVEFs in female young athletes and female veterans is explained by the lower left ventricular diameter and length. A smaller left ventricular diameter and length independently increase LVEF^1, 6^ despite similar strains and a lower wall thickness in the females. The substudy modelling^6^ predicts the young females would have an LVEF 3.1% higher and veteran females 4.9% higher than their male counterparts and close to the 3% and 6% we observed (Appendix Section 5.).

The lower resting blood pressure found in female athletes is consistent with previous observations.^21^ GLS and GCS did not prove useful in distinguishing between the cohorts. Longitudinal stress was, however, more useful in comparing young and veteran athletes but was of no benefit in assessing differences between the sexes.

We found GLASED provided the more reliable assessment of contractance compared with CASED in both this paper using echocardiography and a previous study using CMR.^13^. This may be because circumferential strain varies between ∼-3% to ∼ −35% in the subendocardium and subepicardium respectively.^9^ Furthermore, there is also a large circumferential stress gradient across the wall with very high values subendocardial and low values subepicardial. The combination of heterogenous stresses and strain across the wall may have resulted in the less reliable CASED values. In contrast, the longitudinal stresses and strains are more homogenous resulting in more consistent GLASED results.^13, 24^ Combining GLASED and CASED to give SASED also appeared less useful due to the lower accuracy of CASED.

Reference ranges (95% confidence intervals) varied across our cohorts with values GLASED values less than about 1.5 kJ/m^3^ in young athletes and below 1.0 and 0.9 kJ/m^3^ in veteran males and females respectively. Such reference ranges may be useful in identifying left ventricular myocardial diseases such as hypertrophic and dilated cardiomyopathies where the phenotype is uncertain using conventional findings.

The higher GLASE and longitudinal forces in young male athletes compared to young male non-athletes indicates a training effect brought about by an increase in muscle mass. GLASE provides a measure of the work performed by the whole ventricular muscle mass in the longitudinal direction. GLASE differs from stroke-work as the latter is calculated solely using data from within the lumen alone (i.e. stroke volume and intracavity pressure), while GLASE is calculated with information from myocardium itself. Stroke work does not allow for any changes in ventricular geometry such as increases in wall thickness and ventricular size. GLASE, in contrast, uses information from the myocardium directly by calculating contractile stresses and inputting myocardial strains and, therefore, has theoretical advantages over stroke work. The higher mean GLASE value in males remained greater despite correcting for body size. The variations in GLASE and longitudinal forces may, in part, explain the difference in expected athletes performances in the different age and sex cohorts.

A previous study has assessed GLASED found comparable results. In the CMR study,^13^ the normal cohort had a mean GLASED of 1.94 kJ/m^3^, hypertensive group with early hypertensive cardiomyopathy 1.39 kJ/m^3^, dilated cardiomyopathy 0.86 kJ/m^3^ and amyloid heart disease 0.58 kJ/m^3^. This compares with a GLASED in this echocardiography study of 2.27 kJ/m^3^ and reflects the higher longitudinal shortening (15.4% vs 19.9%). These differences are because the CMR study was based on the long-axis shortening using direct measurements whereas the echocardiogram employed software derived speckle tracking that measured longitudinal strain. Stress was lower in the combine echocardiogram cohort compared to normal controls in the CMR study (22.9 kPa and 25.1 kPa, respectively).

Implementing the calculation of GLASED is straightforward using, for example, the spreadsheet available as a Supplement online which provides the calculation of GLASED using the input variables of systolic blood pressure, left ventricular end-diastolic diameter and end-diastolic wall thickness.

### Limitations

The study utilized GLS measured using software-derived speckle tracking, which may have limitations in terms of accuracy. A precise assessment of contractance is most accurately evaluated using the area under the stress-strain curve, which is readily applicable to ex vivo studies using trabeculae.^12^ However, this numerical integration method is impractical in clinical practice.^13^ An approximation of the area under the stress-strain curve can be made using the more ‘user-friendly’ method derived from the nominal stress. A comparison of GLASED using the simplified equation (analytic method) and longitudinal contractance using numerical integration of the stress-strain curve is presented in the Appendix (Sections 6. and 7.) confirming that the simplified equations use is a reasonable approximation to contractance calculated numerically.

The temporally fluctuating and highly heterogeneous deviatoric and hydrostatic stresses and strains can be expressed with manifold three-dimensional tensors, and the strain energy density calculated from the double dot product of the individual stress and strain tensors (for a linear elastic material 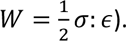. There exists a significant disagreement regarding the values assigned to material properties of myocardium, such as the elastic modulus, which are essential for precise finite element modelling.^25, 26^ The differences in myocardial properties are likely related to the relative health of the tissues undergoing ex vivo analysis with factors such as tissue hypoxia, perfusion, temperature and surrounding milieux. Moreover, the myocardium is hyperelastic and anisotropic, leading to sophisticated modelling that is currently under investigation by our group.^25, 27^ Further, the calculations do not take into account regional dyssynchrony although significant abnormalities would not be expected in these healthy cohorts with normal ECGs. Despite these limitations, the uniaxial approach presented in this study is simple, easy to apply, and, importantly, scalable. Further details supporting the use of nominal rather than instantaneous stress are provided in the Appendix (Sections 6. and 7.). While the estimations of strain energy density may not be perfect using GLASED, it is precise enough for clinical utilization since GLASED performs better than both strain and LVEF in predicting outcome^13, 14^ whilst also reducing the risk of a computational error when compared the numerical method for estimating contractance.

Invasive left ventricular pressures could be used for calculating wall stresses more accurately but were not performed as it is not realistic in clinical practice. Therefore, peak systolic pressure derived noninvasively using a brachial cuff was used as a surrogate. We acknowledge that measuring pressure using a sphygmomanometer may influence the calculation of wall stress. Ambulatory BP results may further improve the accuracy of SBP measurements. Circumferential strain data was not collected the young female group, which prevented the calculation of CASED. We did not have a female control group available, nor did we directly assess the impact of distinct types of sports on GLASED that may have influenced our results. Exercise treadmill testing using VO_2 max_ was not available for these cohorts and so the influence of relative fitness could not be assessed.

Stroke work was not assessed as stroke volume data were not available. Our previous study using MRI,^13, 14^ showed stroke work was unhelpful in predicting mortality. We provided information on myocardial work using GLASE. We suggest that work calculated from luminal data alone (i.e. stroke work) should be improved by incorporating information derived from the actual myocardium using contractance. Regional changes and dyssynchrony were not assessed, however, we do not expect significant regional abnormalities given the healthy cohorts with normal ECGs in this study.

Propagation errors arise when input variables, such as LVIDd, are squared in the GLASED and CASED equations. Therefore, the accuracy of such measurements is crucial for obtaining dependable contractance values. Although we have provided reference ranges for each cohort, we acknowledge that our sample sizes were limited and may not be comparable to studies performed on echocardiographic equipment from different vendors. Nonetheless, we think the reference ranges may be helpful in assessing myocardial function in situations where cardiomyopathic processes are suspected and other measures are inconclusive, as our previous work has shown the clinical utility in disease processes (Appendix Section 5.).^13, 14^

## Conclusions

This observational study utilized echocardiography to assess a novel measure of myocardial contractile function, GLASED. Our findings reveal that young male athletes exhibit higher GLASED values compared to young female athletes, and GLASED decreases with age, while the sex differences observed in young athletes disappear among veteran athletes. Additionally, we explain why there are differences in left ventricular ejection fraction (LVEF) between the sexes.

The results of our study hold significant clinical relevance, as they shed light on myocardial function and its potential implications in the screening of cardiac diseases, including dilated and hypertrophic cardiomyopathies. Specifically, a GLASED value below the reference range may indicate reduced energy production per unit volume of muscle, suggesting the presence of a cardiomyopathic process. This highlights the importance of further investigating the clinical utility of GLASED as a tool for evaluating myocardial function and prognosis in individuals with cardiac disorders.

Our research opens new avenues for understanding and monitoring myocardial function. As such, these findings may contribute to enhanced diagnostic accuracy and improved management of cardiac diseases, particularly in the context of athlete screening and evaluation of borderline phenotypes.

To fully realize the potential benefits of GLASED in clinical practice, future studies should probe deeper into its predictive capabilities and establish standardized reference ranges. The inclusion of GLASED as part of a comprehensive cardiac assessment may lead to improved patient outcomes and better-informed treatment decisions in individuals at risk of or diagnosed with cardiac disorders.

## Data Availability

The corresponding author will share the data underlying this article on reasonable request.

## Appendix

*1. How does LVIDd, wall thickness and myocardial strain impact LVEF?*

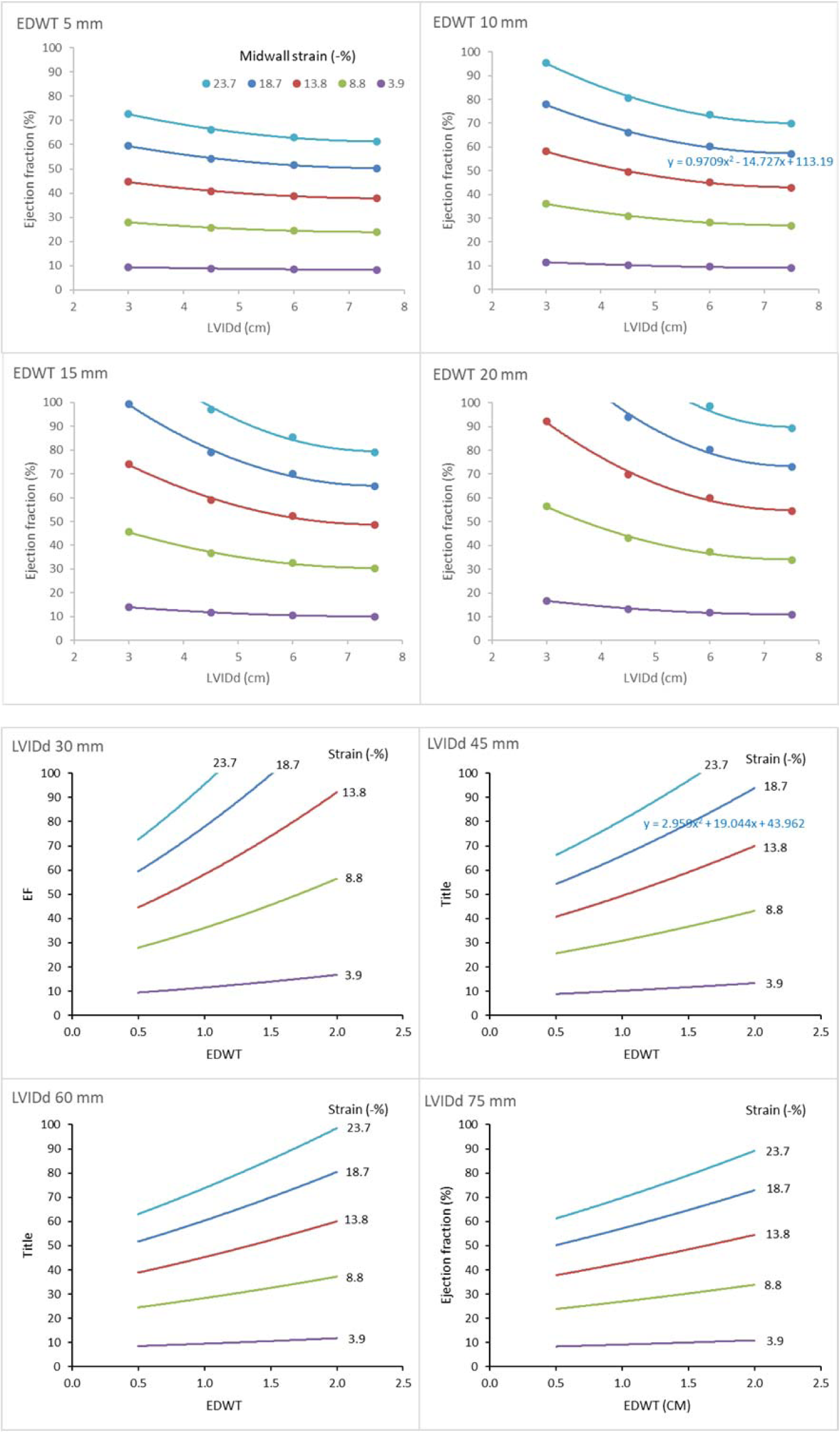
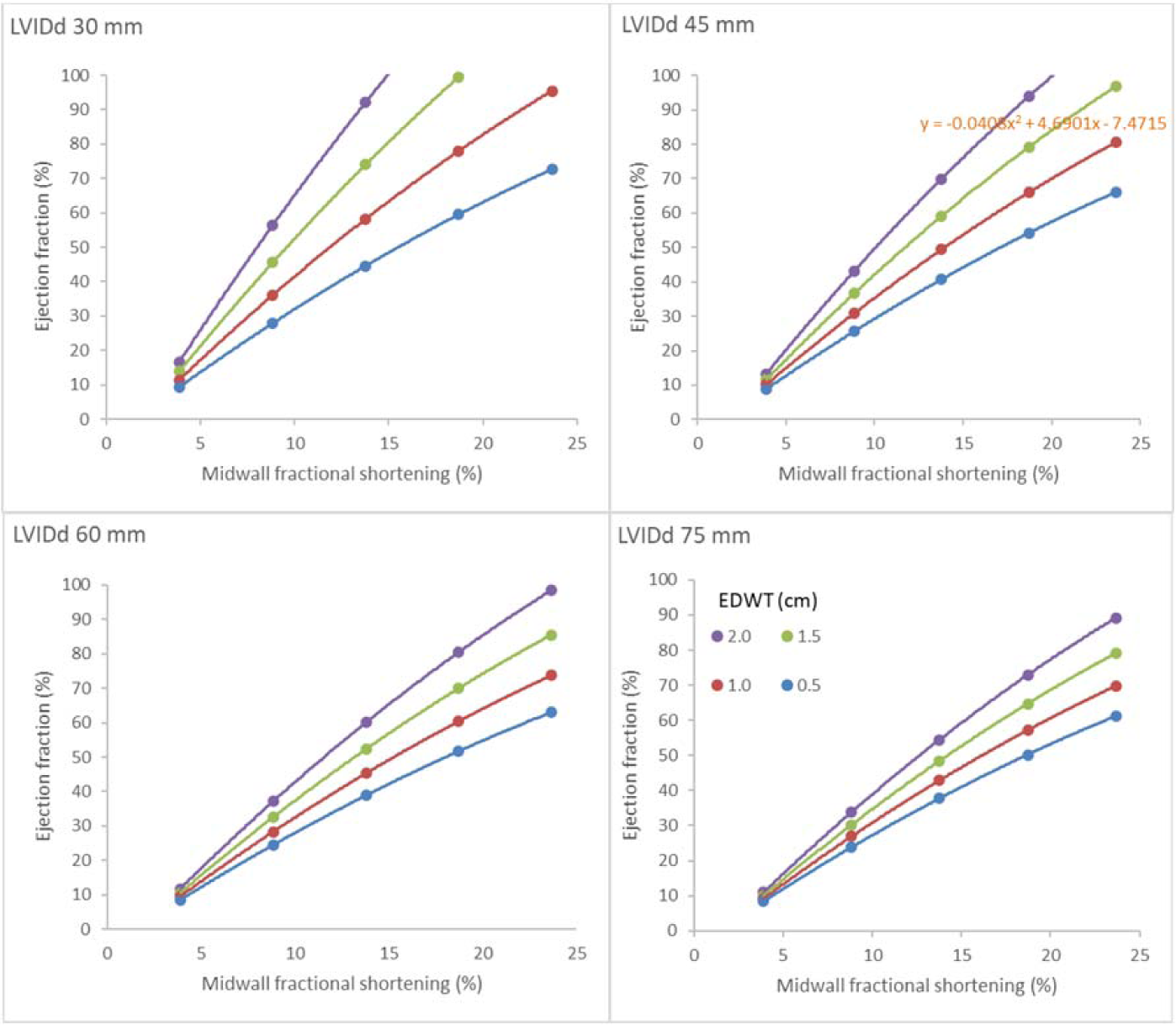

*2. How does GLASED relate to BNP and expected prognosis?*

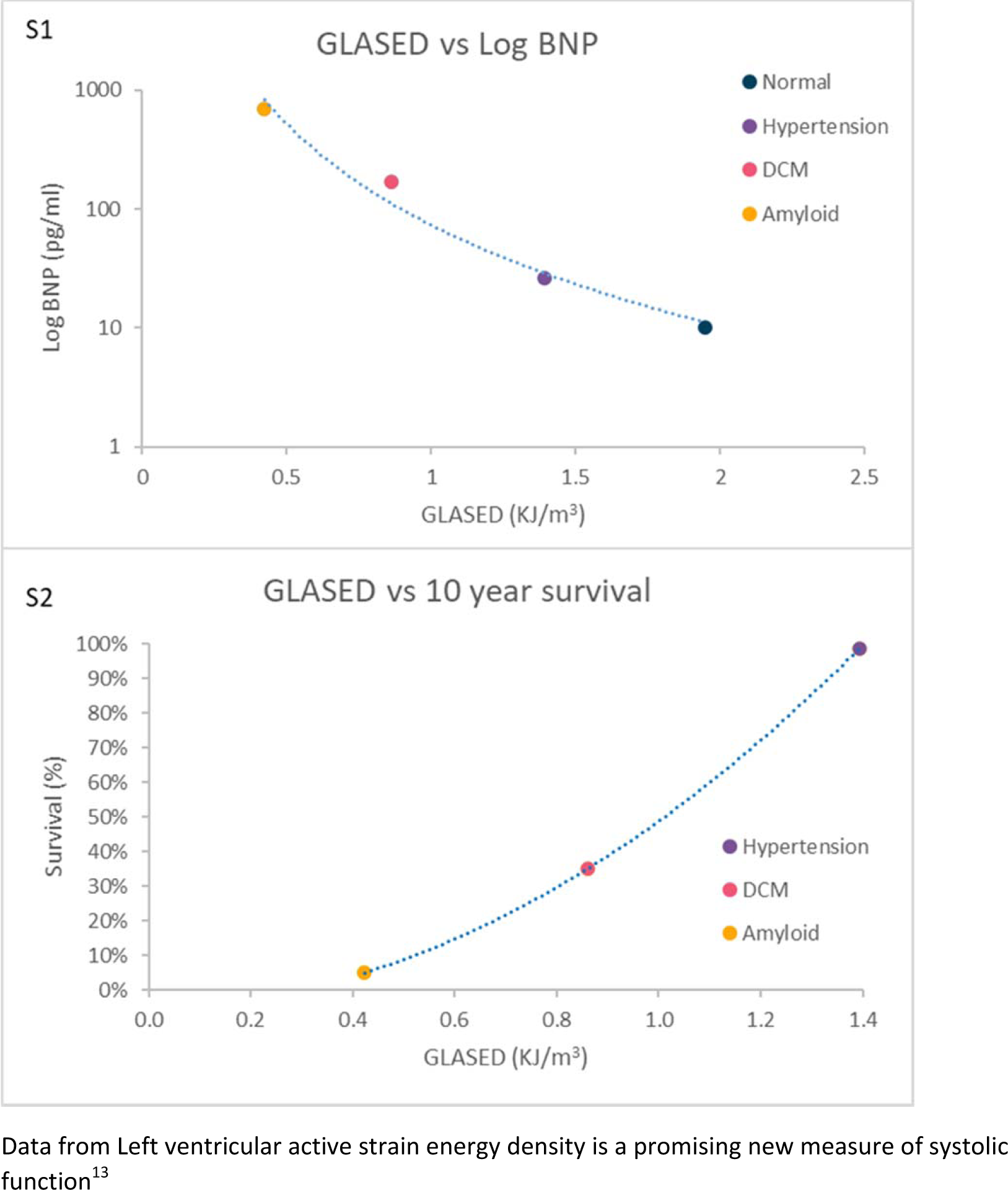

*3. Cox proportional hazard ratios for major adverse cardiovascular events with potential prognostic left ventricular structural and functional markers.*^14^

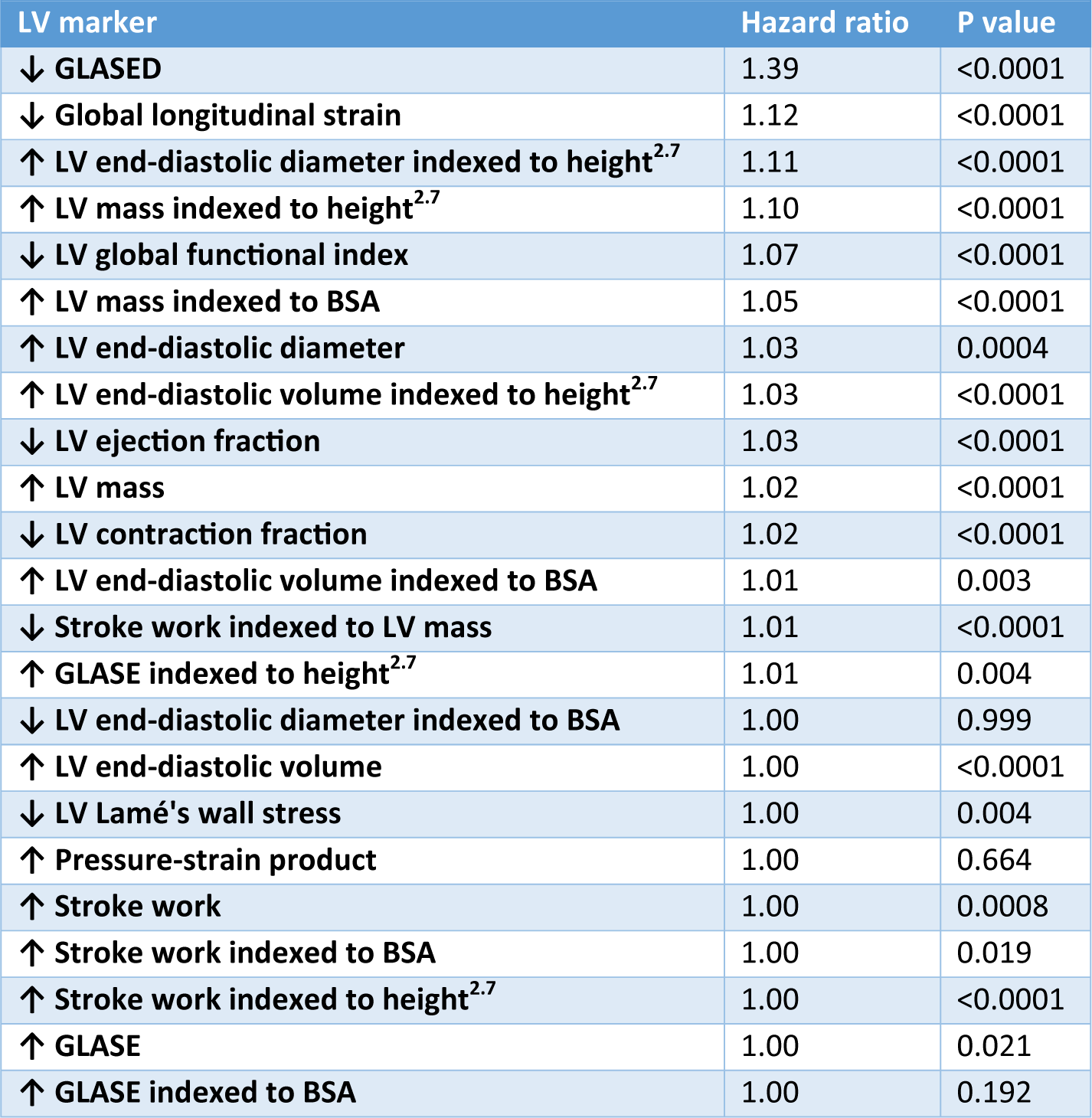

*4. Cox proportional hazard ratios for mortality with potential prognostic left ventricular structural and functional markers.*^14^

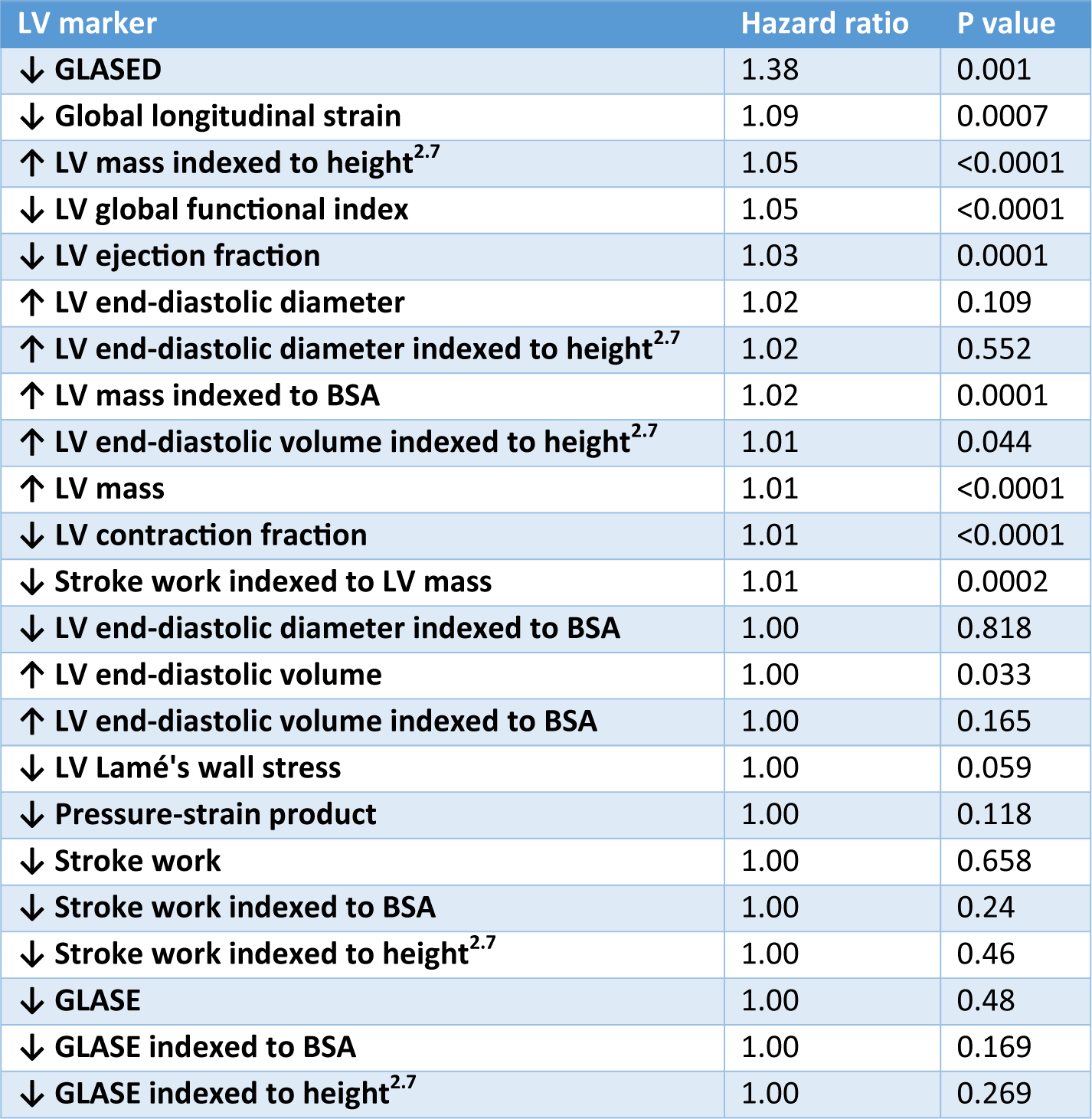

*5. Why is the LVEF higher in females?*

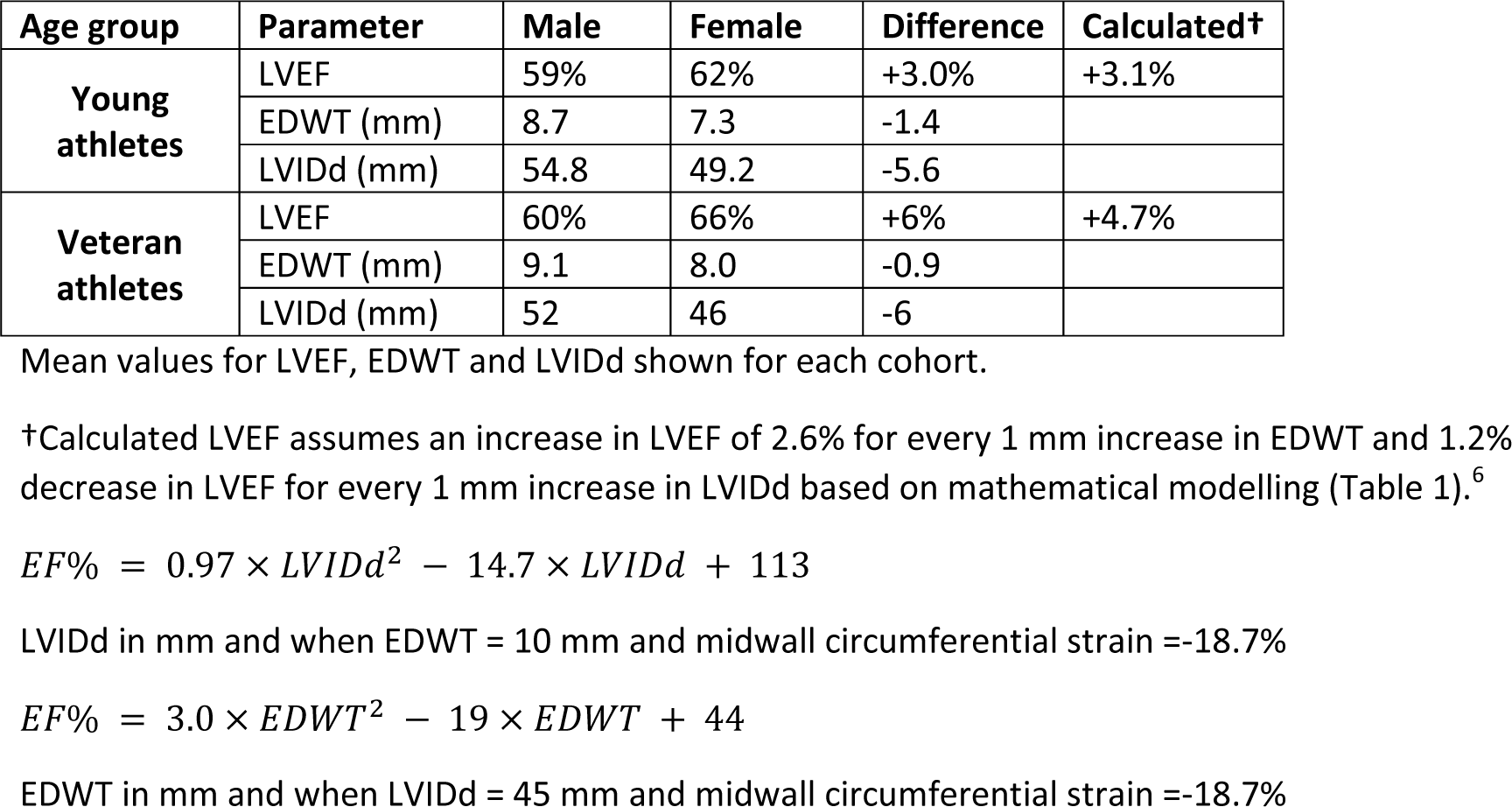

*6. How does longitudinal contractance measured by numerically (using stress-strain curve) vs. analytic method (GLASED equation) compare in normal individuals?*

A typical normal pressure-strain curve was obtained (from Loncaric et al Int J Cardiovasc Imaging 2021 Vol. 37 Issue 1 Pages 145-154) and extracted using Engage digital software. The curve was divided into 12 systole nominal strain intervals (Fig. A). The longitudinal engineering stress was calculated for each strain interval using the Lamé equation to give an approximation of true stress and true strain using the internal dimensions and wall thickness in diastole and systole from a normal group (from D. H. MacIver, et al Sci Rep 2022;12:1-14) (Fig. B).

The active strain energy density was calculated at each change in longitudinal strain and the total obtained by numerical integration using the trapezoidal method giving, in this example, a longitudinal contractance of 2.80 kJ/m^3^ (Fig. C). The active strain energy equates to the area under the blue curve in Figure D. Half the area of the rectangle, peak nominal stress by peak strain (red box) produces the blue triangular area (Fig. D), therefore, the equation [inline1] and results in a GLASED of 2.71 kJ/m^3^ (Fig. 4). The approximation for GLASED is achieved because the vertical hatched area is comparable to the horizontal hatched area. See^13^ for further details.

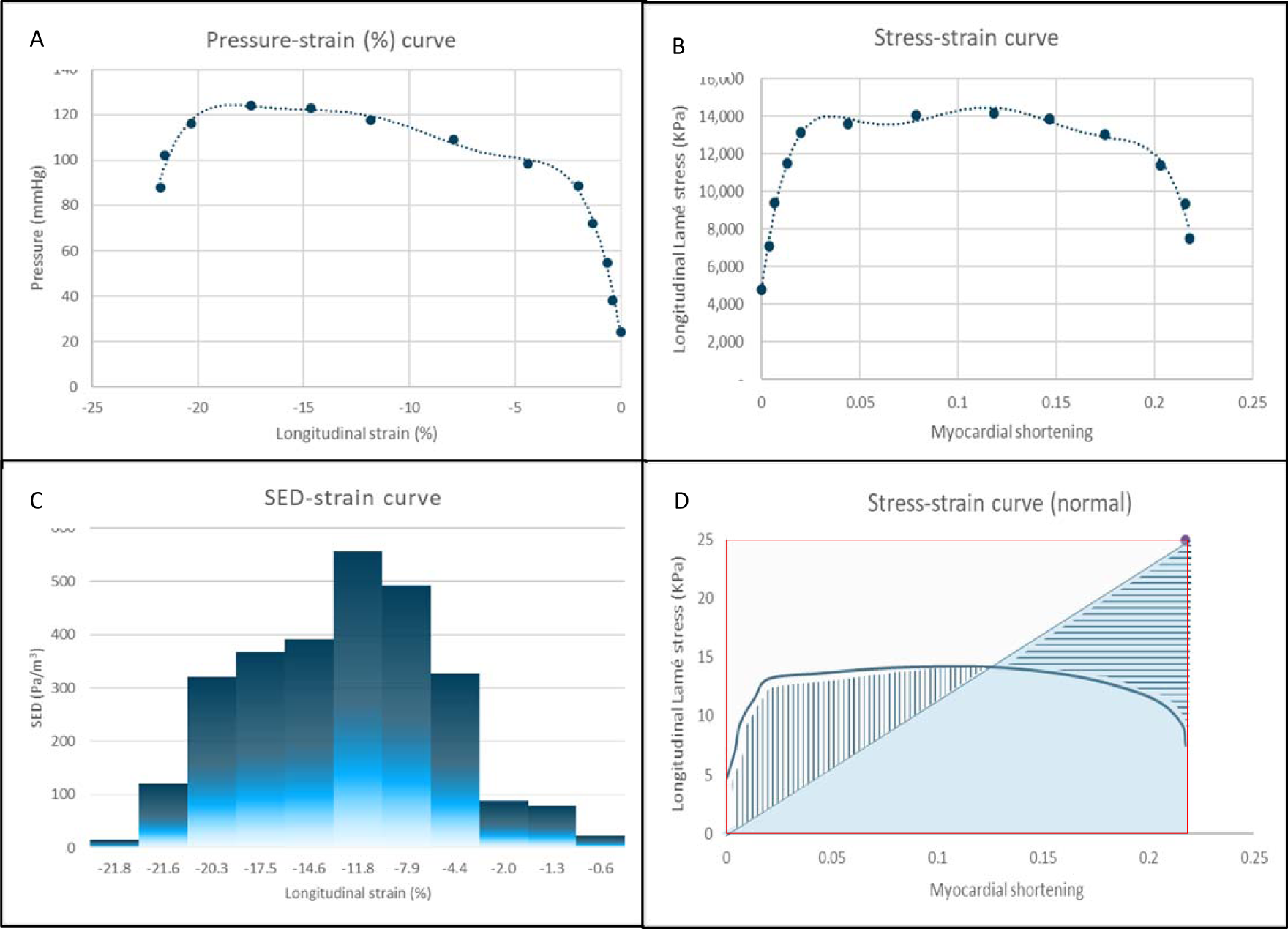

*7. How does longitudinal contractance measured by numerically using stress-strain curve vs. analytic method (GLASED) compare in disease?*

The following data was obtained from “Left ventricular active strain energy density is a promising new measure of systolic function D. H. MacIver, et al Sci Rep 2022;12:1-14” as this provided wide range of GLASED results.

A reference curve was divided into 40 strain points to give 39 intervals and the stress-strain curves plotted as described above for the mean values in the normal controls, hypertensive cardiomyopathy, dilated cardiomyopathy and cardiac amyloid cohorts respectively. Nominal stresses are shown as crosses and true stresses as data points.

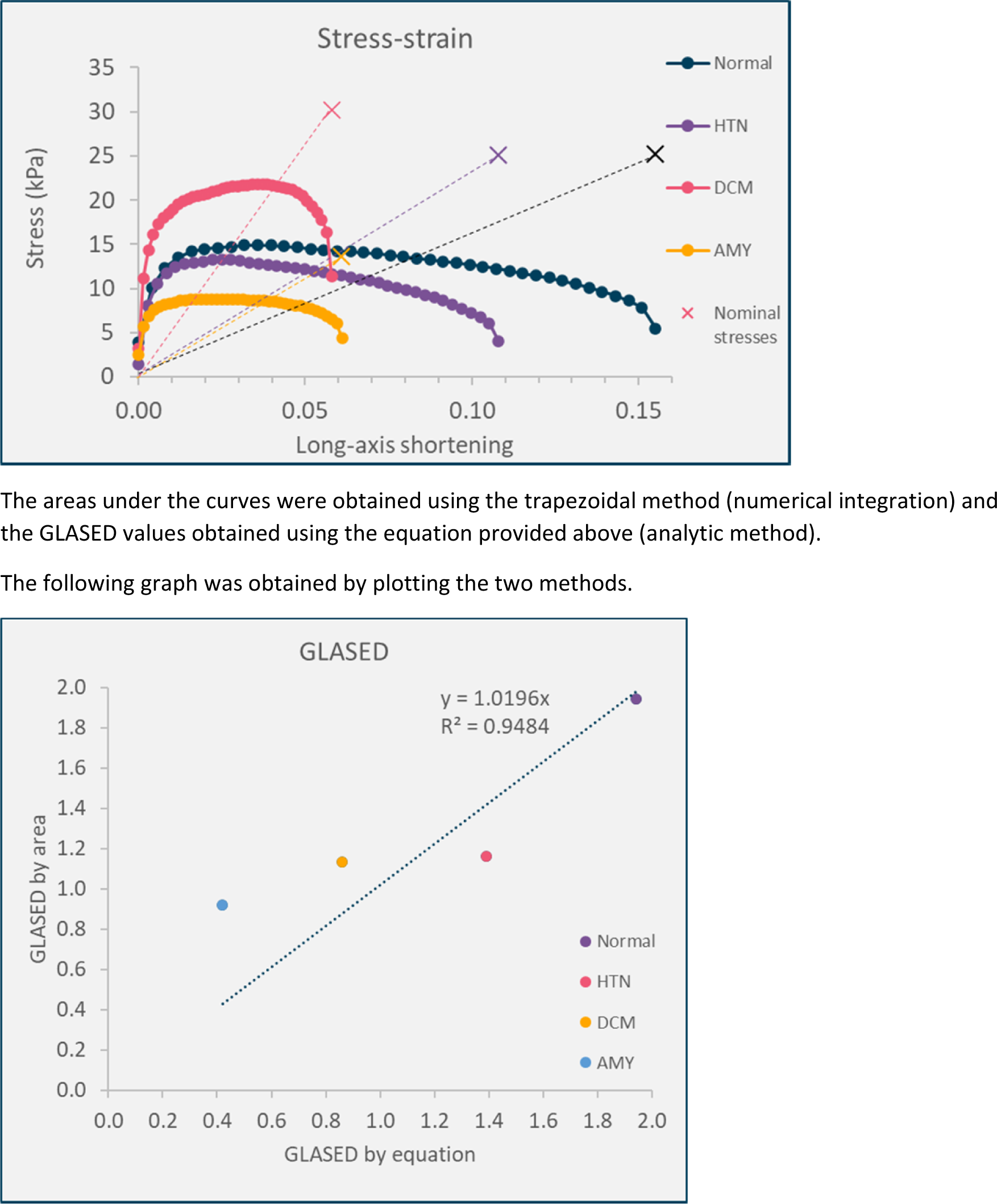

